# A glial-mitochondrial axis in bipolar disorder: in vivo MRI signatures and lithium-associated attenuation

**DOI:** 10.64898/2026.05.30.26354457

**Authors:** Marlene Tahedl, Jonas Rohrer, Sonja Kuster, Ida Mehrdadi, Erich Seifritz, Philipp Homan

**Affiliations:** University Hospital of Psychiatry Zurich, Zurich, Switzerland; University of Zurich, Zurich, Switzerland; Neuroscience Center Zurich, University of Zurich and ETH Zurich, Zurich, Switzerland

**Keywords:** bipolar disorder, glial dysfunction, mitochondrial dysfunction, transcriptomic neuroimaging, lithium treatment, metabolic psychiatry

## Abstract

Bipolar disorder (BD) is associated with widespread white matter microstructural alterations, yet their cellular and metabolic underpinnings remain poorly understood. Here, we asked whether *in vivo* magnetic resonance imaging (MRI) signatures of BD spatially align with the distribution of glial and mitochondrial cell populations, whether these patterns are specific to BD across the affective-psychotic spectrum, and whether lithium attenuates them. In individuals with BD (*n* = 104), major depressive disorder (MDD; *n* = 135), and psychotic disorders (PY; *n* = 87) from the UK Biobank, each matched to healthy controls, we mapped multimodal MRI alterations (radial diffusivity [RD], fractional anisotropy [FA], voxel-based morphometry [VBM]) onto reference maps of five glial cell types and six mitochondrial markers. BD showed a reproducible spatial alignment between elevated radial diffusivity and glial-rich regions (astrocytes, microglia, endothelial cells, oligodendrocyte precursors), together with a separable alignment between regional gray-matter loss and mitochondrial respiratory capacity. Across diagnostic groups, psychotic disorders partially shared the glial signature but lacked the mitochondrial one, while MDD diverged on both, supporting a degree of BD specificity for the combined glial-mitochondrial pattern. Within BD, lithium-treated patients showed an attenuation of glial alignment most prominently for astrocytes and oligodendrocyte precursors, consistent with a glial mechanism of lithium action. While effect magnitudes were modest, as is typical for cross-modal spatial alignment studies, they were consistent across markers and modalities. The findings identify glial-mitochondrial coupling as a tractable cellular axis in BD pathophysiology and point to glial pathways as a candidate substrate for lithium’s therapeutic effect.

## Introduction

Bipolar disorder (BD) is a severe, chronic psychiatric condition characterized by recurrent episodes of mania and depression, affecting approximately 1–2% of the population worldwide and ranking among the leading causes of disability [1, 2]. Diffusion MRI consistently reveals abnormal white matter microstructure in BD, with the most reproducible finding being increased radial diffusivity across major commissural and association tracts including the corpus callosum, cingulum, and superior longitudinal fasciculus [3–5]. Because radial diffusivity is biologically interpreted as a marker of myelin integrity and the glial cells that support it, this pattern points toward a glial or myelin-related substrate. Findings for fractional anisotropy in BD are more heterogeneous in direction and anatomical location [4–6], and gray matter volume reductions detected by voxel-based morphometry [7–9], though present, are less consistent across studies. What remains unclear is which cellular populations these specific microstructural and volumetric changes reflect, and whether the cellular basis of BD is distinct from that of related affective and psychotic disorders.

Convergent evidence implicates glial dysfunction and mitochondrial bioenergetic impairment in mood disorders, particularly in BD [10–12]. Glial cells (astrocytes, microglia, oligodendrocyte precursor cells) and mitochondria jointly regulate brain energy metabolism, synaptic support, inflammation, and oxidative stress, which are processes repeatedly implicated in BD [13, 14]. These metabolic pathways are themselves tightly linked to circadian regulation, which is frequently disrupted in BD and shows in vivo neuroanatomical correlates in hypothalamic structure that vary by chronotype [15], potentially contributing to the observed bioenergetic deficits. However, most supporting evidence derives from post-mortem, peripheral biomarker, or preclinical studies. *In vivo* spatial mapping that directly links MRI alterations to regional glial and mitochondrial marker expression in the living human brain has remained scarce.

Lithium remains one of the most effective and best-studied treatments for BD, particularly for preventing manic episodes and reducing suicide risk [16, 17]. Preclinical and emerging clinical data indicate that lithium acts on glial cells, especially astrocytes and oligodendrocyte precursor cells, and modulates mitochondrial function and energy metabolism [18–22]. Yet, direct *in vivo* evidence linking lithium treatment to regional glial or mitochondrial signatures in the human brain is still limited. Comparing lithium-treated and lithium-untreated BD individuals within a spatial-alignment framework can identify the cellular populations whose MRI signatures are associated with lithium exposure, providing a within-disorder probe of treatment-relevant cellular targets that complements between-disorder specificity analyses. A transdiagnostic comparison addresses a related question. Affective and psychotic disorders show substantial clinical and biological overlap, and traditional categorical diagnoses have limited neurobiological validity [23, 24]. Comparing BD, major depressive disorder (MDD), and psychotic disorders (PY) within the same analytic framework allows direct testing of whether glial and mitochondrial signatures are BD-specific, shared across the affective-psychotic spectrum, or follow a graded pattern across diagnoses. Such designs align with dimensional frameworks [23, 24] and turn diagnostic specificity from an assumption into an empirically testable outcome.

The present study addressed these questions in sequence. Using the *JuSpace* toolbox [25] in a large, well-characterized UK Biobank (UKB) sample, we first asked whether MRI signatures in BD (RD, FA, VBM) align spatially with reference maps of glial and mitochondrial cell populations. We then tested whether any such alignment is specific to BD relative to matched MDD and PY groups, or shared across the affective-psychotic spectrum. Finally, within BD, we asked whether lithium treatment is associated with attenuation of the identified signatures, providing a within-disorder cellular probe of treatment effects. Effect sizes in cross-modal spatial-alignment studies are typically modest. We therefore emphasize consistency across markers and modalities, together with diagnostic specificity, as the primary criteria for interpreting the findings.

## Methods and Materials

### Participants

The present study utilized multimodal neuroimaging data (T1w and dMRI) from the UK Biobank (UKB, [26]), a large-scale population-based resource. From the 90,088 participants with both T1w and dMRI data available at baseline, diagnostic groups were first identified using linked hospital and primary-care ICD-10 codes. The primary group of interest was BD (ICD-10 code F31x, *n* = 113 initial). HC had no recorded psychiatric ICD-10 diagnoses (*n* = 9,798 eligible). **Figure 1** shows a flow chart of participant selection.

**Figure 1.**
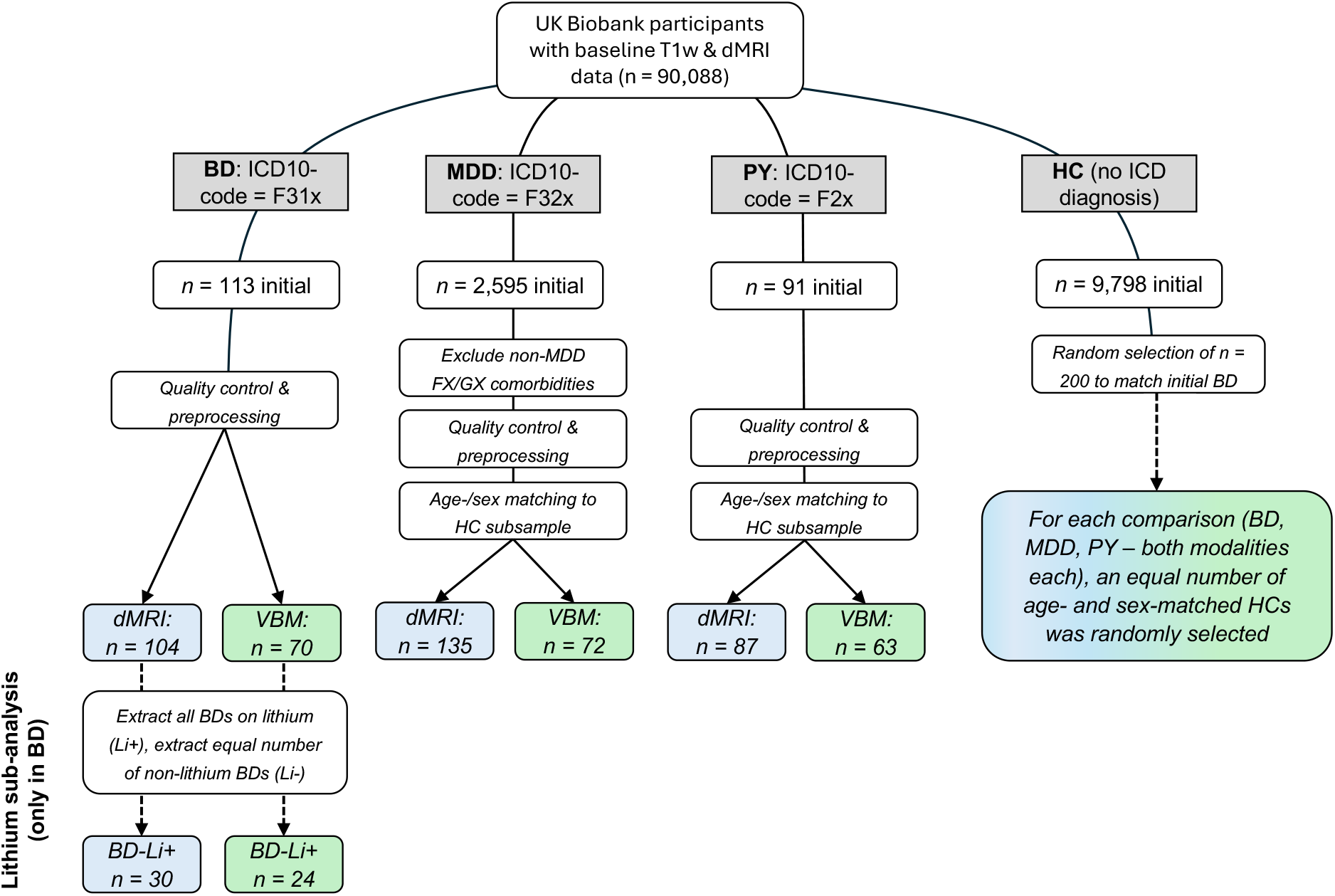
Flowchart illustrating participant selection from the UK Biobank. Diagnostic groups were identified using ICD-10 codes. For each comparison, an equal number of age- and sex-matched HCs was randomly selected from all eligible HC participants. Lithium sub-analysis was conducted within BD patients only. Abbreviations: BD = bipolar disorder; dMRI = diffusion-weighted magnetic resonance imaging; ICD = international classification of diseases; Li(+/–) = lihtium (taking/non-taking); MDD = major depressive disorder; PY = psychotic disorders; T1w = T1-weighted; VBM = voxel-based morphometry

To facilitate efficient matching and prepare for subsequent transdiagnostic analyses (particularly the larger MDD cohort), a random subsample of *n* = 200 HC participants was first drawn from the eligible pool. This step reduced computational demands while maintaining a sufficiently large reservoir from which age- and sex-matched controls could be selected for all diagnostic groups. For the BD versus HC comparison, HC participants were randomly subselected from this *n* = 200 pool and matched to the BD group on age and sex. Because the *JuSpace* toolbox requires equal sample sizes, the final matched groups were further reduced (still preserving age and sex balance) to *n* = 104 for diffusion MRI and *n* = 70 for voxel-based morphometry (VBM) after quality control and preprocessing.

For the lithium sub-analysis within BD, all available BD patients taking lithium were included (dMRI: *n* = 30 Li+; VBM: *n* = 24 Li+). An equal number of age- and sex-matched lithium-untreated BD patients (Li−) was then randomly selected from the remaining BD participants to form the comparator groups.

Transdiagnostic specificity analyses followed the same logic: From the full MDD pool (F32x/F33x, *n* = 2,595 initially with multimodal neuroimaging data), participants were subsampled, quality-controlled, and preprocessed, yielding final groups of *n* = 135 (dMRI) and *n* = 72 (VBM). Age- and sex-matched HC were drawn from the *n* = 200 pool for each modality. The same procedure was applied to the PY disorders group (F2x, *n* = 91 initially), resulting in final samples of *n* = 87 (diffusion MRI) and *n* = 63 (VBM) with corresponding matched HC. Note that final sample sizes differed between dMRI and VBM modalities because of varying data availability in the UKB neuroimaging repository and additional quality-control exclusions during preprocessing. All patient groups were pre-matched to HC on age and sex for each modality separately. Handedness distributions and total intracranial volume were comparable across groups (see Table 1), which was confirmed using unpaired *t*-tests (for age and TIV) and Chi-squared tests (for sex and handedness distributions) within *Matlab* (version R2024b).

**Table 1.**
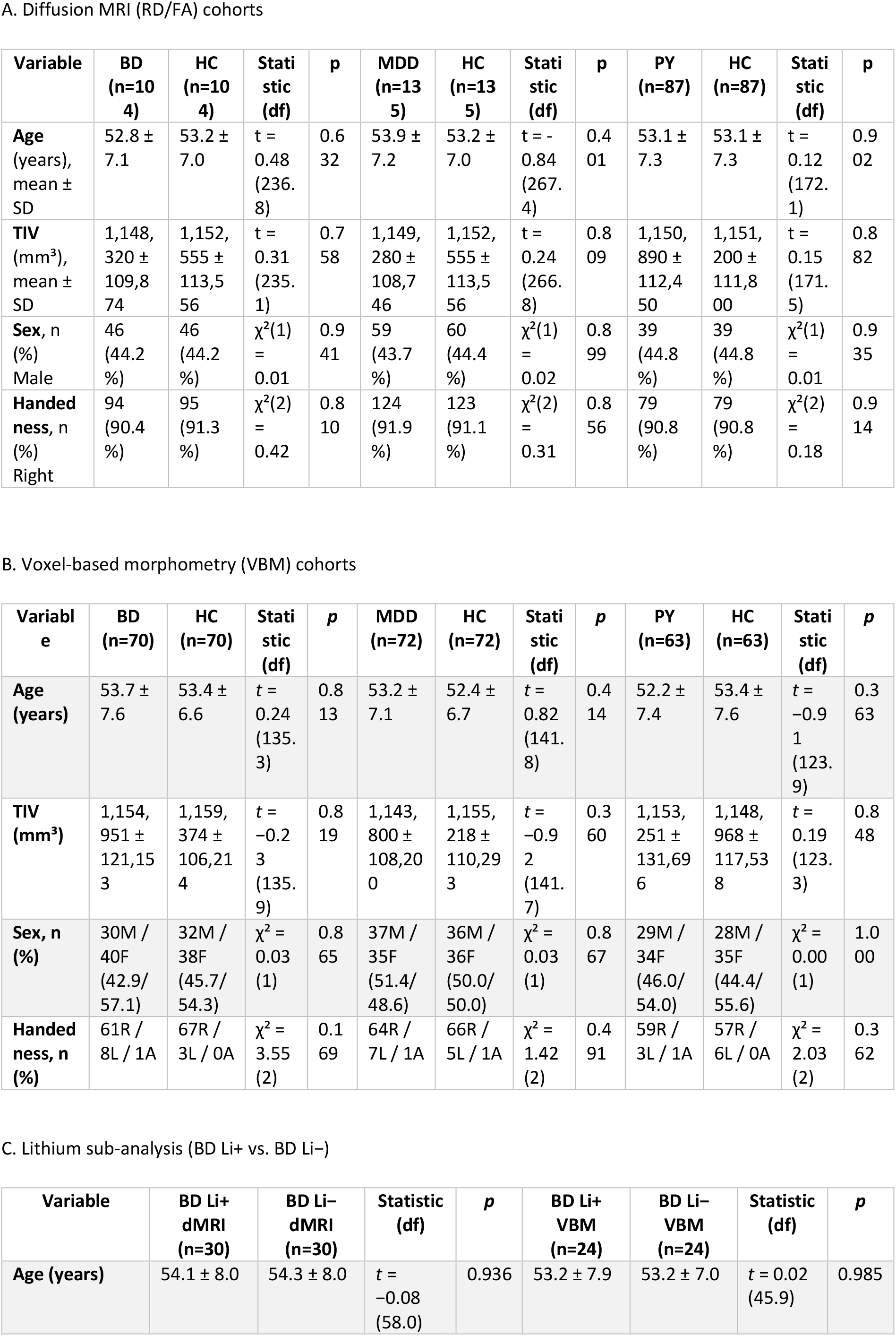
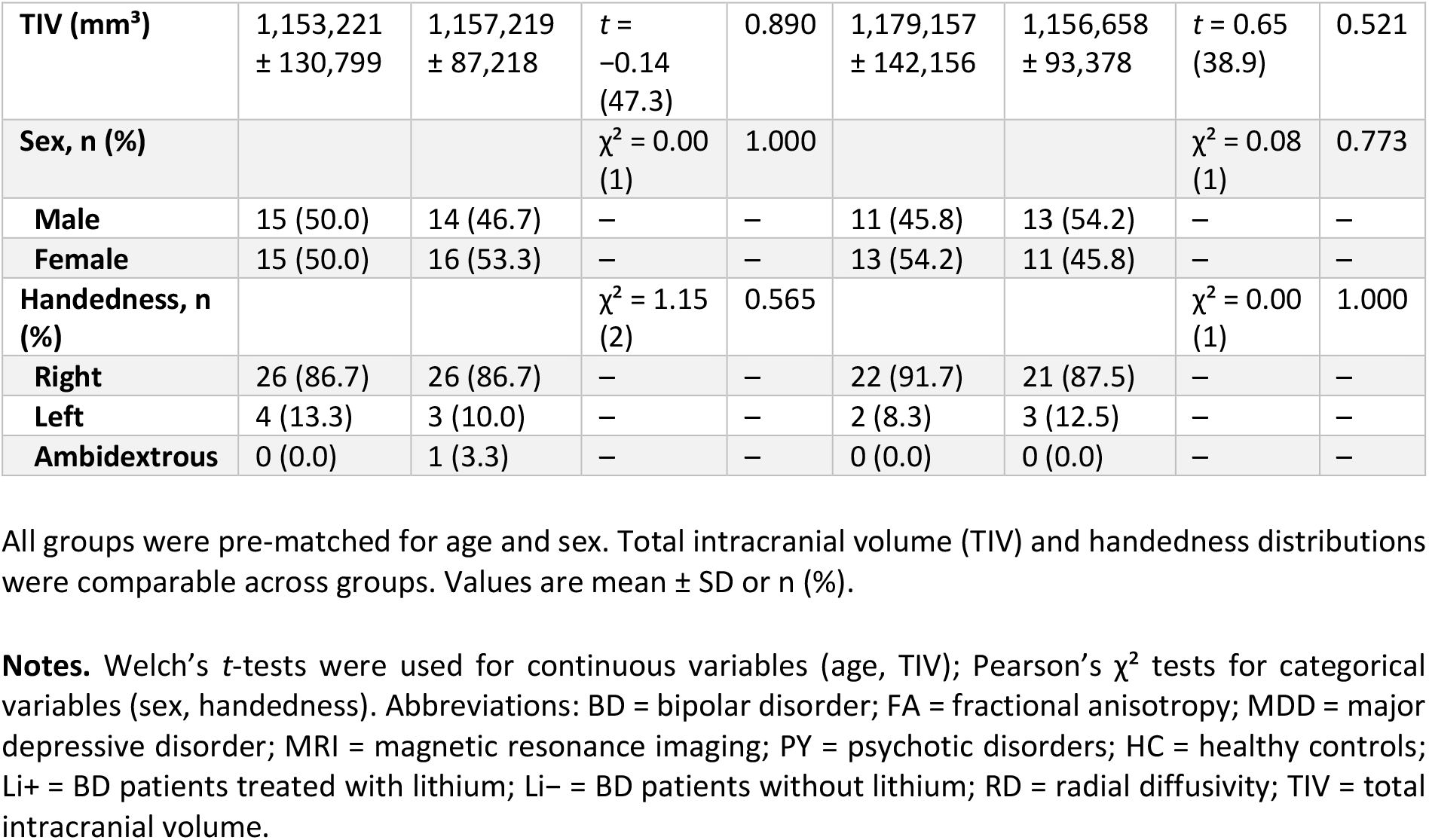
Demographic characteristics of the study samples.

UKB has an ethics approval from the North West Multi-centre Research Ethics Committee (reference 11/NW/0382) and all participants gave written informed consent. Our study was conducted under the UKB Application Number 102266.

### Neuroimaging data acquisition and preprocessing

All MRI data were acquired as part of the UKB brain imaging protocol on 3 Tesla Siemens Skyra scanners using a 32-channel head coil. The T1w structural images were obtained with a 3D MPRAGE sequence (1 mm isotropic resolution, field of view 208 × 256 × 256 mm, acquisition time ~5 min). dMRI images were acquired with a multi-shell protocol (2 mm isotropic resolution, 100 diffusion directions across *b*=1000 and *b*=2000 s/mm^2^shells plus *b*=0 images, acquisition time ~7 min) ([27]; see also UK Biobank Brain Imaging Documentation at https://biobank.ndph.ox.ac.uk/showcase/showcase/docs/brain_mri.pdf).

T1w images were processed using FSL’s [28, 29] VBM pipeline ([30], https://fsl.fmrib.ox.ac.uk/fsl/docs/structural/fslvbm.html) with standard options except for the omission of spatial smoothing, yielding gray and white matter concentration maps. The procedure was described in detail earlier [31]. dMRI data underwent comprehensive preprocessing adapted from a recently published state-of-the-art protocol optimised for UKB data [32]. All steps, including denoising, Gibbs-ringing correction, motion and eddy-current correction, susceptibility-induced distortion correction, and bias-field correction, were performed exclusively using *MRtrix3* tools [33] on a Linux computing cluster. From the fully preprocessed dMRI data in individual space, the diffusion tensor was fitted and FA and RD maps were derived. For downstream preprocessing within *JuSpace* (cf. below), the FA/RD maps were non-linearily warped into MNI152 standard space (2mm resolution). All resulting maps underwent rigorous quality control before inclusion in subsequent analyses.

### JuSpace correlational and statistical analysis

For each subject, an individual MRI difference map was computed by subtracting the group-mean HC map (matched modality and sample) voxel-wise from the subject’s individual map. Each subject-level difference map was then spatially correlated (Spearman’s ρ) with each cellular marker map, yielding one ρ per subject per marker. Group differences were tested by permutation of group labels (5,000 permutations), with significance assessed against the resulting null distribution and FDR correction applied across markers within each contrast. These spatial correlations were performed using the *Matlab*-based *JuSpace* toolbox [25]. The toolbox (version 2.1, downloaded from https://github.com/juryxy/JuSpace on 14 May 2026) integrates MRI data with nuclear imaging-derived neurotransmitter maps as well as cellular marker maps. The latter include five glial cell-type maps (astrocytes, endothelial cells, microglia, oligodendrocyte precursor cells [OPC], and mature oligodendrocytes) derived from the Human Brain Atlas (AHBA, https://brain-map.org, processed via the *abagen* toolbox; [34– 36]) and six mitochondrial maps (Complex1-NADH, Complex2-SDH, Complex4-COX, RespiratoryCapacity, TissueDensity, TissueRespCapacity) based on a recent high-resolution human brain map of mitochondrial respiratory capacity and diversity, also based on AHBA data [13].

Consistent with our study objectives – identifying BD-specific pathologic signatures linked to brain metabolism and circadian regulation – we restricted analyses to the glial and mitochondrial cellular marker maps (11 maps in total). The AHBA-derived maps are based on post-mortem tissue from six donors and show a known cortical bias (most samples originate from cortical regions, with limited subcortical and white-matter coverage), but they remain the current gold-standard reference for spatially resolved cellular expression in the human brain.

For each diagnostic contrast (BD vs. HC, MDD vs. HC, PY vs. HC) and the lithium sub-analysis (BD Li+ vs. Li−), voxel-wise Spearman rank correlations were computed between the MRI group-difference maps (RD, FA, and VBM) and each cellular marker map. Directionality of contrasts was consistently set as patient group > HC (or Li+ > Li−) unless otherwise specified. Spearman correlations were chosen because they are non-parametric and robust to non-normal distributions and outliers common in spatial neuroimaging data, as outlined in the original *JuSpace* publication [25] (cf. also **Fig. 2**). Because *JuSpace* currently supports only pairwise comparisons with equal sample sizes and does not allow inclusion of covariates, all groups were pre-matched on age and sex, and sample sizes were balanced accordingly for each modality (cf. section “Participants”). Partial-volume correction was applied using T1w tissue probability maps to account for tissue mixing at the voxel level. Significance was assessed with 5,000 permutations, and resulting *p*-values were corrected for multiple comparisons using the false-discovery-rate (FDR) procedure [37]. We report FDR-corrected *p*-values (*p*_FDR_) together with the median Spearman’s ρ for each marker.

**Figure 2.**
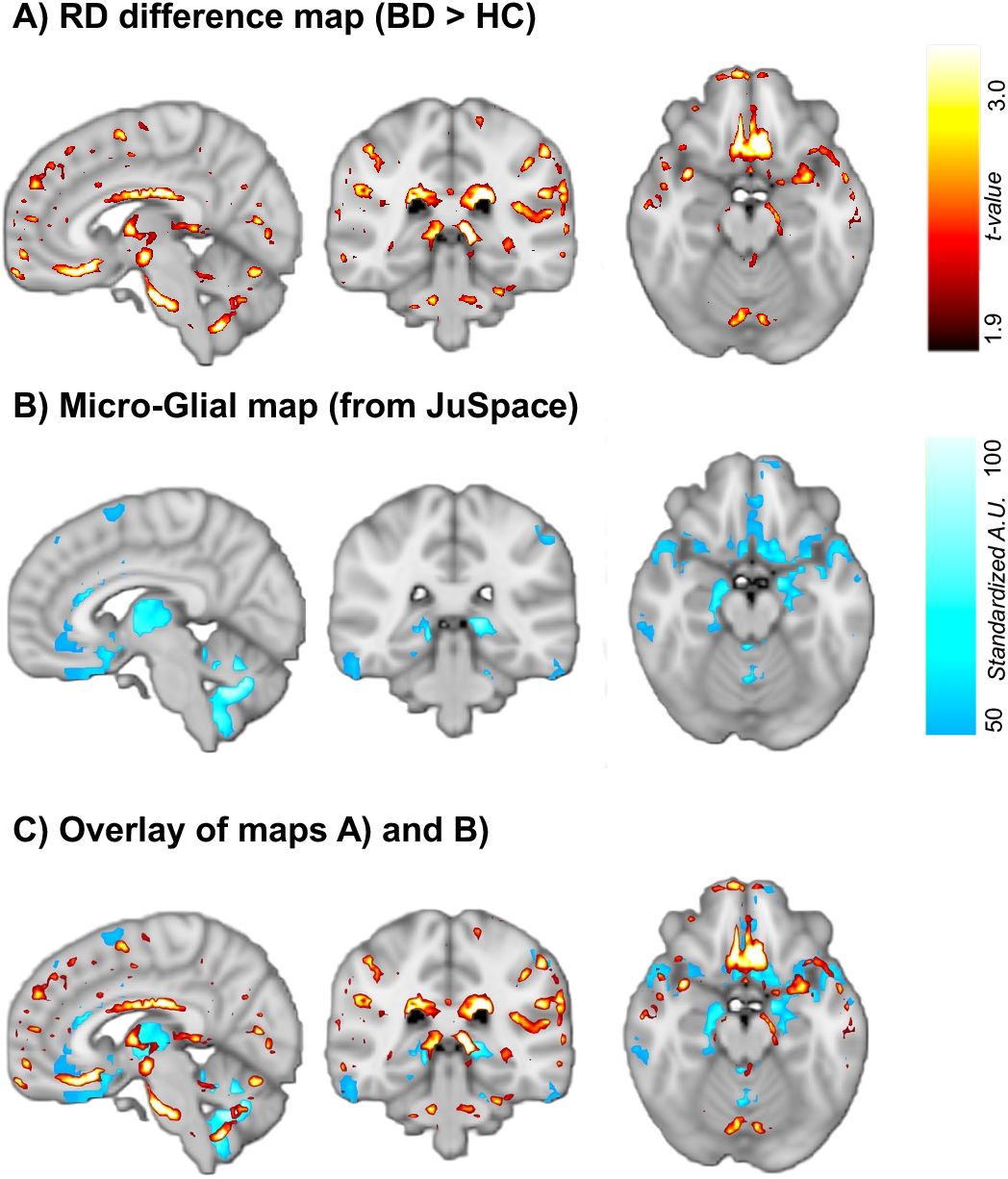
Example of spatial correspondence between neuroimaging metrics and cellular marker expression in BD. (A) Statistical *t*-map showing regions with increased RD in BD compared to HCs (calculated using FSL’s ‘randomise’ tool, 5,000 permutations). (B) Microglia density map derived from the Allen Human Brain Atlas (provided within the JuSpace toolbox; standardized arbitrary units). (C) Overlay of the RD *t*-map shown in (A) and the microglia density map shown in (B), illustrating their spatial alignment. We used JuSpace to perform voxel-wise spatial Spearman correlations (5,000 permutations, FDR-corrected) between group-difference neuroimaging maps and regional cellular marker expression. Abbreviations: BD = bipolar disorder; HC = healthy controls; RD = radial diffusivity; TFCE = Threshold-Free Cluster Enhancement; FSL = FMRIB Software Library

## Results

### Participants characteristics

Final analysed samples after quality control, preprocessing, and matching comprised 104 BD, 135 MDD, and 87 PY participants, each compared against equal numbers of age- and sex-matched HC for dMRI, and 70 BD, 72 MDD, and 63 PY participants with matched HC for VBM (see **Fig. 1** for selection flowchart). A lithium sub-analysis was performed within BD (dMRI: *n* = 30 Li+ vs. *n* = 30 Li−; VBM: *n* = 24 Li+ vs. *n* = 24 Li−). Age and sex matching was successful across all comparisons (cf. Tab. 1). TIV and handedness distributions were comparable between patient groups and HC, with no significant differences (all *p* > 0.16, cf. **Tab. 1**).

### BD versus HC

In the primary analysis, BD was associated with a robust spatial pattern of increased RD that significantly correlated with glial cellular marker expression (**Fig. 3A, Tab. 2**). Specifically, higher RD in BD vs HC overlapped with regions of high expression of astrocytes (ρ = 0.058, *p*_FDR_ = 0.021), endothelial cells (ρ = 0.041, *p*_FDR_ = 0.028), microglia (ρ = 0.078, *p*_FDR_ = 0.002), and OPC (ρ = 0.093, *p*_FDR_ = 0.002). Oligodendrocytes showed only a trend (*p*_FDR_ = 0.082). No significant correlations were observed with mitochondrial markers.

**Figure 3.**
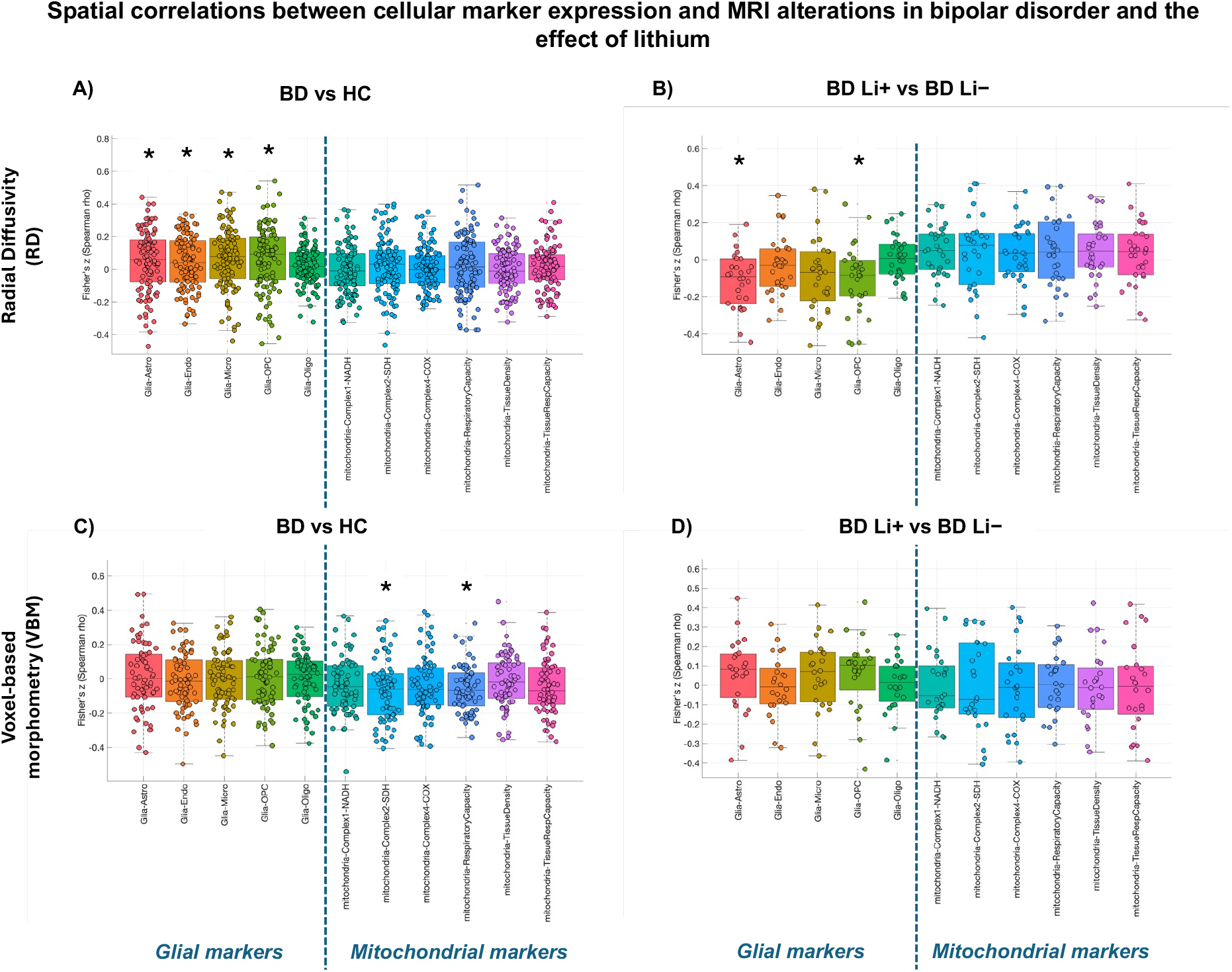
JuSpace spatial correlation results showing cellular marker expression aligned with MRI alterations in BD (left pannels) and the modulatory effect of lithium (right panels). (A, B) Results for RD. (C, D) Results for VBM. Left panels (A, C) show BD vs HC, right panels (B, D) show the lithium sub-analysis (BD Li+ vs BD Li−). Glial markers are shown on the left and mitochondrial markers on the right of each panel. Values represent median Spearman’s ρ. Asterisks indicate significant correlations after FDR correction (*p*_FDR_ < 0.05). Abbreviations: BD = bipolar disorder; HC = healthy controls; RD = radial diffusivity; VBM = voxel-based morphometry; Li+ = lithium-treated; Li− = lithium-untreated; FDR = false discovery rate

**Table 2.** Spatial correlations between cellular marker expression maps and MRI-based measures of cerebral pathology in affective and psychotic disorders.

| Cellular marker | BD vs HC<br>[median<br>Spearman's $\rho$,<br>($p_{FDR}$)] | BD Li+ vs Li-<br>[median<br>Spearman's $\rho$,<br>($p_{FDR}$)] | MDD vs HC<br>[median<br>Spearman's $\rho$,<br>($p_{FDR}$)] | PY vs HC [median<br>Spearman's $\rho$,<br>($p_{FDR}$)] |
| --- | --- | --- | --- | --- |
| **Diffusion-weighted MRI-derived: Radial diffusivity (RD)** |  |  |  |  |
| **Glial** |  |  |  |  |
| Astro | 0.058 (**0.021**) | -0.090 (**0.020**) | 0.024 (0.286) | 0.040 (**0.037**) |
| Endo | 0.041 (**0.028**) | -0.030 (0.357) | 0.014 (0.715) | 0.064 (**0.037**) |
| Micro | 0.078 (**0.002**) | -0.068 (0.064) | -0.010 (0.534) | 0.067 (**0.037**) |
| OPC | 0.093 (**0.002**) | -0.085 (**0.049**) | 0.022 (0.286) | 0.061 (**0.037**) |
| Oligo | 0.019 (0.082) | 0.009 (0.842) | 0.003 (0.722) | 0.027 (0.373) |
| **Mitochondrial** |  |  |  |  |
| Complex1-NADH | -0.009 (0.831) | 0.052 (0.128) | 0.012 (0.174) | -0.014 (0.867) |
| Complex2-SDH | 0.032 (0.381) | 0.076 (0.345) | 0.017 (0.286) | 0.013 (0.732) |
| Complex4-COX | -0.002 (0.687) | 0.033 (0.345) | 0.016 (**0.043**) | -0.005 (0.732) |
| RespiratoryCapacity | 0.015 (0.429) | 0.042 (0.345) | 0.014 (0.286) | -0.007 (0.732) |
| TissueDensity | -0.011 (0.934) | 0.047 (0.175) | 0.021 (**0.043**) | -0.009 (0.867) |
| TissueRespCapacity | 0.019 (0.443) | 0.044 (0.345) | 0.035 (0.078) | -0.012 (0.732) |
| **Diffusion-weighted MRI-derived: Fractional anisotropy (FA)** |  |  |  |  |
| **Glial** |  |  |  |  |
| Astro | -0.009 (0.576) | 0.002 (0.984) | -0.005 (0.967) | -0.019 (0.724) |
| Endo | -0.044 (0.279) | 0.017 (0.984) | -0.011 (0.967) | -0.060 (0.649) |
| Micro | -0.009 (0.606) | 0.025 (0.984) | 0.005 (0.967) | -0.036 (0.649) |
| OPC | -0.017 (0.606) | 0.027 (0.984) | 0.003 (0.967) | -0.017 (0.649) |
| Oligo | -0.032 (0.194) | 0.022 (0.984) | -0.021 (0.967) | -0.002 (0.649) |
| **Mitochondrial** |  |  |  |  |
| Complex1-NADH | -0.005 (0.364) | -0.049 (0.984) | -0.021 (0.967) | 0.008 (0.888) |
| Complex2-SDH | -0.011 (0.606) | 0.006 (0.984) | -0.014 (0.967) | -0.005 (0.888) |
| Complex4-COX | -0.014 (0.606) | -0.019 (0.984) | -0.005 (0.967) | 0.004 (0.937) |
| RespiratoryCapacity | -0.011 (0.712) | -0.027 (0.984) | 0.004 (0.967) | 0.002 (0.888) |
| TissueDensity | -0.011 (0.262) | -0.036 (0.984) | -0.015 (0.967) | -0.009 (0.888) |
| TissueRespCapacity | -0.009 (0.621) | 0.002 (0.984) | -0.002 (0.967) | 0.012 (0.888) |
| **T1-weighted MRI-derived: voxel-based morphometry (VBM)** |  |  |  |  |
| **Glial** |  |  |  |  |
| Astro | 0.003 (0.704) | 0.082 (0.679) | -0.024 (0.268) | -0.017 (0.712) |
| Endo | -0.015 (0.704) | -0.008 (0.954) | -0.007 (0.268) | 0.024 (0.712) |
| Micro | 0.002 (0.974) | 0.071 (0.679) | -0.041 (0.268) | 0.004 (0.712) |
| OPC | 0.013 (0.704) | 0.104 (0.679) | -0.050 (0.313) | -0.021 (0.712) |
| Oligo | 0.026 (0.974) | 0.015 (0.954) | 0.016 (0.922) | 0.022 (0.912) |
| **Mitochondrial** |  |  |  |  |
| Complex1-NADH | -0.045 (0.145) | -0.054 (0.954) | 0.043 (0.268) | 0.023 (0.712) |
| Complex2-SDH | -0.058 (**0.007**) | -0.072 (0.954) | 0.033 (0.538) | -0.037 (0.712) |
| Complex4-COX | -0.064 (0.145) | -0.011 (0.954) | 0.011 (0.268) | 0.029 (0.712) |
| RespiratoryCapacity | -0.068 (**0.004**) | 0.004 (0.954) | 0.012 (0.837) | -0.017 (0.769) |
| TissueDensity | -0.019 (0.704) | -0.012 (0.954) | 0.056 (0.121) | 0.027 (0.712) |
| TissueRespCapacity | -0.071 (0.077) | -0.006 (0.954) | 0.028 (0.268) | 0.008 (0.913) |

Values are presented as median Spearman's $\rho$ ( $p_{FDR}$ ). Bold indicates FDR-corrected $p < 0.05$ . All values rounded to three significant digits.
Abbreviations: BD = bipolar disorder; FDR = false discovery rate; MDD = major depressive disorder; PY = psychotic disorders; HC = healthy controls; Li+ = lithium-treated; Li- = lithium-untreated. Analyses were performed within *JuSpace* using 5,000 permutations, partial volume correction, and using a whole-brain mask. For further details on the methods, cf. also Methods section of the main text.

In contrast, VBM analyses revealed a different pattern (**Figs. 3C–D, Tab. 2**). BD was associated with significantly negative spatial correlations between GM volume and mitochondrial markers, most prominently Complex2-SDH (ρ = −0.058, *p*_FDR_ = 0.007) and RespiratoryCapacity (ρ = −0.068, *p*_FDR_ = 0.004). No significant correlations were observed with glial markers.

No significant spatial correlations were found for FA in any contrast (all *p*_FDR_ > 0.19; see **Supplementary Fig. S1**).

### Diagnostic specificity

To evaluate the specificity of the observed patterns to BD, we conducted parallel analyses in MDD and PY. **Figure 4** shows the corresponding analyses in MDD and PY, for direct comparison with the BD pattern (and the effects of lithium, see below) in Figure 3.

**Figure 4.**
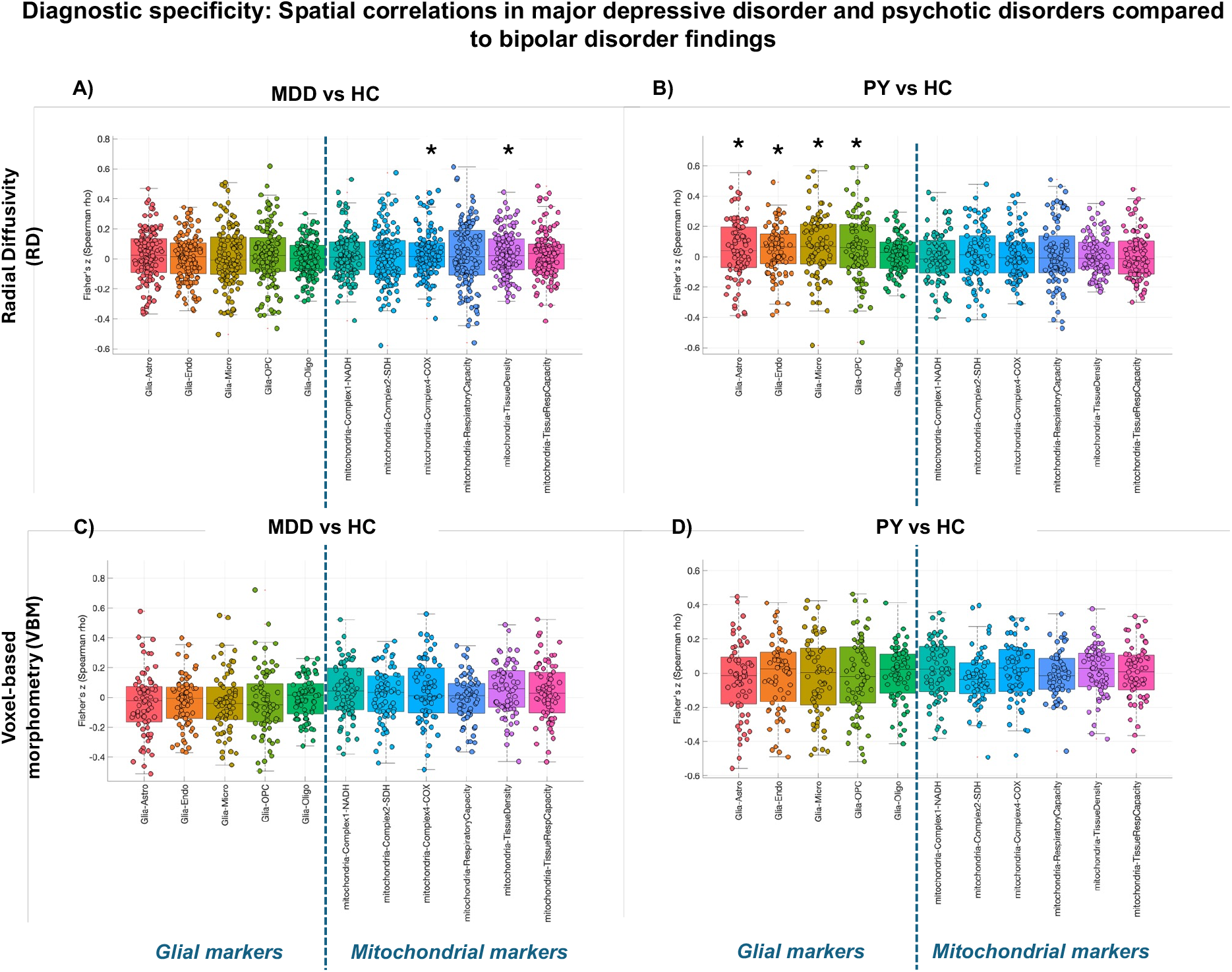
Diagnostic specificity of spatial correlations between cellular marker expression and MRI alterations. (A, B) Results for RD. (C, D) Results for VBM. Left panels show major MDD vs HC, right panels show PY vs HC. Glial markers are displayed on the left and mitochondrial markers on the right of each panel. Values represent median Spearman’s ρ. Asterisks indicate significant correlations after FDR correction (*p*_FDR_ < 0.05). Together with Figure 3, these results highlight a relatively specific glial-RD signature in BD (and partially in PY) and a mitochondrial-VBM signature primarily in BD. Abbreviations: BD = bipolar disorder; MDD = major depressive disorder; PY = psychotic disorders; HC = healthy controls; RD = radial diffusivity; VBM = voxel-based morphometry; FDR = false discovery rate

In MDD vs HC, the glial-RD signature observed in BD was absent (**Fig. 4A, Tab. 2**). Instead, RD alterations showed significant positive spatial correlations with mitochondrial markers, particularly Complex4-COX ρ = 0.016, (*p*_FDR_ = 0.043) and TissueDensity (ρ = 0.021, *p*_FDR_ = 0.043), with a trend for TissueRespCapacity (ρ = 0.035, *p*_FDR_ = 0.078). No significant correlations emerged for glial markers or in the VBM analysis (all *p*_FDR_ > 0.12; **Fig. 4C, Tab. 2**).

In PY vs HC, results more closely resembled the BD profile for RD (**Fig. 4B, Tab. 2**). Significant positive correlations were found with glial markers, including astrocytes (ρ = 0.040, *p*_FDR_ = 0.037), endothelial cells (ρ = 0.064, *p*_FDR_ = 0.037), microglia (ρ = 0.067, *p*_FDR_ = 0.037), and OPC (ρ = 0.061, *p*_FDR_ = 0.037). However, unlike BD, no significant mitochondrial-VBM correlations were observed in PY (all *p*_FDR_ > 0.71; **Fig. 4D, Tab. 2**).

FA again showed no significant spatial correlations in either MDD or PY (all *p*_FDR_ > 0.19; **Supplementary Fig. S1**).

### Effects of lithium in bipolar disorder

This glial-RD signature observed in BD vs. HC. was partially reversed by lithium treatment (**Fig. 3B, Tab. 2**). In the lithium sub-analysis (BD Li+ vs. Li−), we found significant negative correlations with astrocytic (ρ = −0.090, *p*_FDR_ = 0.020) and OPC markers (ρ = −0.085, *p*_FDR_ = 0.049), with a trend for microglia (ρ = −0.068, *p*_FDR_ = 0.064). This indicates that lithium treatment was associated with lower RD in regions of high glial expression. No significant spatial correlations were found for FA in any contrast (all *p*_FDR_ > 0.19; see **Supplementary Fig. S1**). Moreover, the lithium sub-analysis did not yield significant effects on VBM-mitochondrial correlations.

## Discussion

The present study identified two distinct cellular signatures of bipolar disorder in vivo: a spatial alignment between elevated radial diffusivity and glial-rich regions, and a separable alignment between regional gray-matter volume loss and mitochondrial respiratory capacity. The glial signature was partially shared by psychotic disorders, while major depressive disorder diverged from both patterns, indicating relative (though not absolute) specificity of the combined signature to BD. Within BD, lithium-treated individuals showed an attenuation of the glial signature most prominently for astrocytes and oligodendrocyte precursors, consistent with a glial mechanism of lithium action; the mitochondrial signature was not modulated by lithium status. FA showed no spatial alignment in any contrast. Effect magnitudes were modest, as is expected for cross-modal spatial alignment studies in psychiatric disorders, but were consistent across markers and across MRI modalities and align with biologically interpretable directions. Together, the findings position glial and mitochondrial substrates as two partially dissociable axes of BD pathophysiology and identify glia as a candidate mediator of lithium’s clinical effects.

The spatial alignment between elevated RD and regions of high glial expression is consistent with glial involvement in the white matter pathology of BD. RD is biologically interpreted as a marker of myelin integrity, with secondary sensitivity to the astrocytic and oligodendroglial populations that maintain it [3, 38]. An alignment between RD elevations and the regional distribution of glial cell types is therefore the spatial pattern one would expect if BD-related microstructural change were preferentially affecting glia-rich territory. The convergence across four glial cell types (astrocytes, microglia, endothelial cells, oligodendrocyte precursors) is consistent with the broader literature implicating multiple glial populations in BD: postmortem studies have reported reduced astrocytic and oligodendroglial densities in prefrontal cortex [39–41], altered microglial morphology and inflammatory signaling in subsets of patients, e.g. those with a history of suicide [42–44], and astrocytic metabolic dysfunction in patient-derived iPSCs [45]. Whether the alignment reflects primary glial pathology, secondary glial response to neuronal injury, or a combination of both cannot be resolved from cross-sectional spatial data; what the present findings establish is that the regional distribution of BD-related microstructural change is anatomically organized in a way that tracks glial cell-type density, providing in vivo anatomical evidence consistent with a causal role for glial populations in BD white matter pathology.

The transdiagnostic comparisons reveal a partially graded pattern rather than a clean hierarchy. The glial-RD signature was present in BD and partially shared by PY (significant alignment with the same four glial cell types, though with a more compressed effect distribution), while MDD showed no glial-RD alignment. The mitochondrial-VBM signature, by contrast, was specific to BD. Neither MDD nor PY showed any significant alignment between gray-matter loss and mitochondrial reference maps. This partial-overlap structure fits the data better than the linear “affective-psychotic spectrum” ordering sometimes invoked in dimensional frameworks [23, 24]. One interpretation is that microstructural change in glia-rich territory may be shared across disorders characterised by recurrent severe affective or psychotic episodes (BD and PY), while cumulative bioenergetic strain sufficient to produce a detectable mitochondrial-aligned volume signature may be more characteristic of BD. The reported MDD mitochondrial-RD alignment was very small in magnitude (median ρ < 0.02) and should be regarded as exploratory rather than as evidence of a divergent mechanism.

Within BD, the glial signature was attenuated in lithium-treated patients, most prominently for astrocytic and oligodendrocyte precursor markers. The effect-size shift (medians of approximately −0.10 ρ units) was visibly larger than in the BD versus HC comparison, making the lithium analysis the most quantitatively pronounced finding in the present study despite its smaller sample. Two interpretations are compatible: a direct cellular effect of lithium on the astrocytic and OPC populations whose markers track the BD signature, consistent with experimental evidence on lithium’s glial targets [45, 46]; or a selection effect, in which lithium-treated patients differ from untreated patients on unmodelled dimensions (lithium responsiveness, illness chronicity, episode polarity, concurrent medication) that drive both prescription and attenuated alignment. A cross-sectional design cannot adjudicate between these, and we therefore cannot claim that lithium causally rescues glial pathology – only that the spatial signature differs systematically with treatment status in directions consistent with the experimental literature. The mitochondrial-VBM signature showed no parallel lithium effect, suggesting the two BD signatures are dissociable in their relationship to treatment as well as in their cellular substrates.

These findings illustrate both the potential and the limits of cross-modal spatial alignment for studying cellular pathology in psychiatric disorders in vivo. Where post-mortem studies measure cells directly but outside the brain context and in vivo MRI captures anatomy but is silent on cellular composition, spatial alignment offers an intermediate inference: whether disease-related anatomical change is spatially organised in ways that track known cellular distributions. The recovered magnitudes are correspondingly modest. This a feature of the design, and the contribution lies in the convergent pattern across markers, modalities, and diagnostic groups rather than in any single ρ value. Three lines of follow-up would meaningfully advance this work: independent replication in cohorts with deeper clinical phenotyping than UKB provides, spatial-autocorrelation-preserving null models (such as variogram-matched surrogates) to test robustness under more conservative inference, and longitudinal designs with within-subject lithium initiation to convert the cross-sectional lithium contrast into a causal test. Until then, the present findings should be read as in vivo anatomical evidence consistent with a glial-mitochondrial axis of BD pathophysiology.

Several limitations should be noted. First, the JuSpace permutation null shufles group labels but does not preserve spatial autocorrelation in the cellular reference maps; spatial-null-robust frameworks were not available within the toolbox version used here, and replication under variogram-matched surrogates remains a needed sensitivity analysis. Second, the cellular reference maps are derived from post-mortem tissue with known cortical bias and limited subcortical and white-matter coverage, which makes the interpretation of RD findings in cortical territory and of VBM findings in white-matter territory indirect [38, 47, 48]. Third, UKB psychiatric diagnoses rely on linked hospital and primary-care ICD-10 codes with acceptable but imperfect validity and likely under-represent milder cases [49, 50]. Fourth, JuSpace does not currently allow covariate modelling, so medication beyond lithium, illness duration, BMI, and other metabolic variables could not be statistically controlled. The lithium sub-analysis provided a direct within-BD contrast but, as noted above, cannot itself establish causality. Finally, sample sizes were moderate, particularly for VBM and the lithium contrast, reflecting the trade-off between matching stringency and power.

In sum, this study identifies two partially dissociable cellular signatures of BD in vivo: a glial-RD alignment partially shared with psychotic disorders, and a mitochondrial-VBM alignment relatively specific to BD. Together with a within-disorder lithium contrast, this points to glia as a candidate mediator of treatment effects. The magnitudes are modest and the inference is anatomical rather than causal, but the convergence across markers, modalities, and diagnostic groups suggests that spatial alignment between in vivo MRI and reference cellular atlases is a tractable approach to mechanism-informed psychiatric neuroimaging.

**Supplementary Figure S1.**
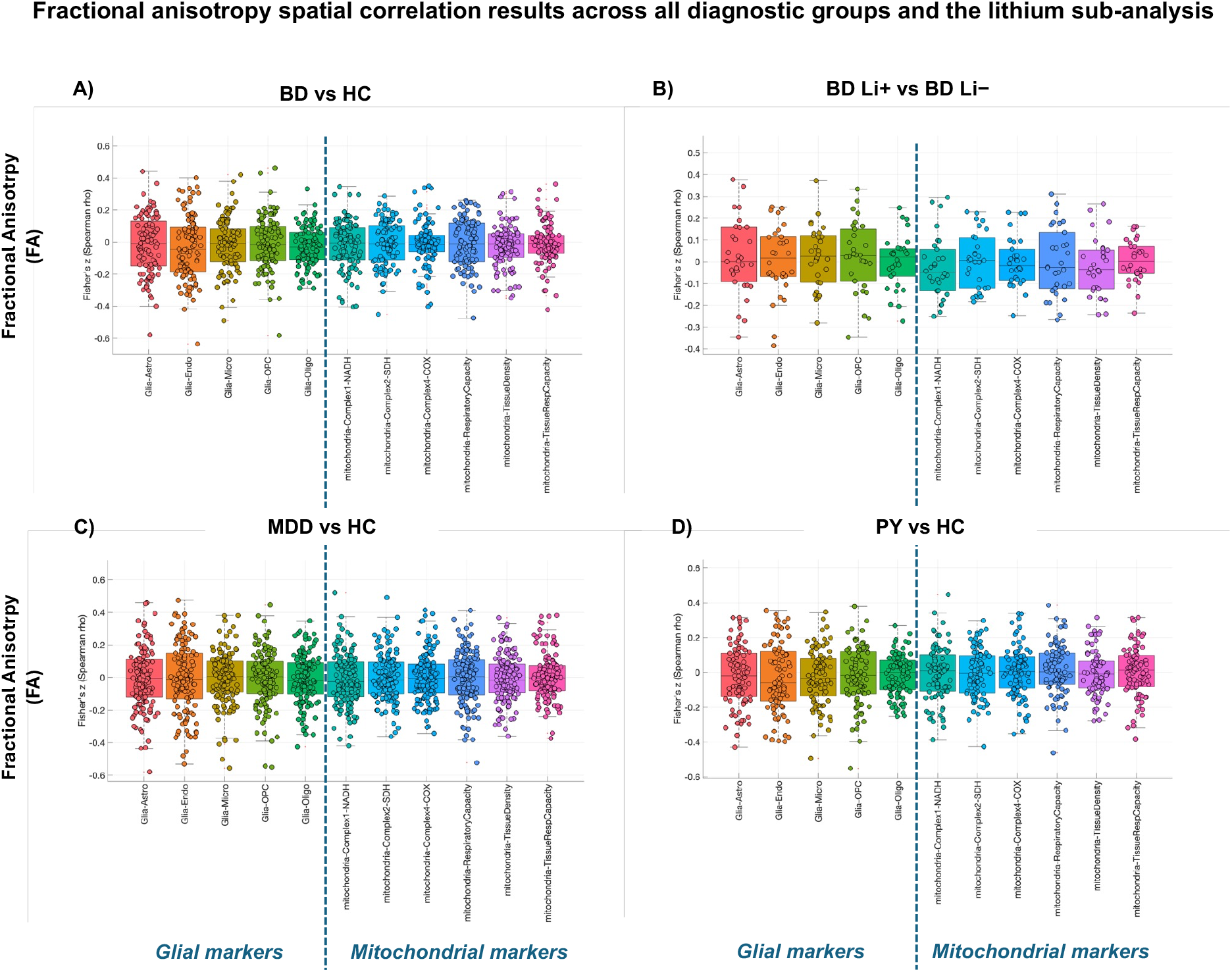
FA spatial correlation results. (A, B) Results for BD vs HC and the lithium sub-analysis (BD Li+ versus BD Li−). (C, D) Results for MDD vs HC and PY vs HC. Glial markers are shown on the left and mitochondrial markers on the right of each panel. Values represent median Spearman’s ρ. No correlations reached statistical significance after FDR correction (all *p*_FDR_ > 0.19). Layout and conventions follow main Figures 3 and 4. These results demonstrate the relative specificity of RD over FA for detecting glial and metabolic signatures. Abbreviations: BD = bipolar disorder; HC = healthy controls; MDD = major depressive disorder; PY = psychotic disorders; Li+ = lithium-treated; Li− = lithium-untreated; FDR = false discovery rate

## CRediT authorship contribution statement

**MT:** Conceptualization, Data curation, Formal analysis, Methodology, Visualization, Writing – original draft, Writing – review & editing. **JR:** Formal analysis, Validation, Writing – review & editing. **SK:** Validation, Writing – review & editing. **IM:** Validation, Writing – review & editing. **ES**: Funding acquisition, Writing – review & editing. **PH:** Conceptualization, Funding acquisition, Supervision, Writing – review & editing.

## Acknowledgments

MT is supported by a project grant from the Hans und Marianne Schwyn-Stiftung and by a Filling-the-Gap fellowship of the Faculty of Medicine, University of Zurich.

## Disclosures

PH has received grants and honoraria from Novartis, Lundbeck, Mepha, Janssen, Boehringer Ingelheim, Neurolite, and OM Pharma outside of this work. No other disclosures were reported. ES received honoraria from Lundbeck Switzerland, Lundbeck Denmark and Switzerland, OM Pharma Switzerland, Recordati Switzerland, Otsuka Switzerland, Mepha Pharma Switzerland, Schwabe Pharma Switzerland and Germany, all were unrlated to this work.

## Data availability

The data used in this study were obtained from the UK Biobank under approved application access. UK Biobank is a large-scale biomedical database that provides controlled access to its data for qualified researchers, subject to application and approval. The data are not publicly available but can be accessed through the UK Biobank repository by researchers who meet the access criteria. The analysis scripts used in this study are available from the corresponding author upon reasonable request.

## Notes

**Conflicts of interest statement:** PH has received grants and honoraria from Novartis, Lundbeck, Mepha, Janssen, Boehringer Ingelheim, Neurolite, and OM Pharma outside of this work. All other authors report no biomedical financial interests or potential conflicts of interest.

### Author Declarations

UK Biobank (https://ams.ukbiobank.ac.uk/ams/)

